# Automated monitoring of movement disorders in dyskinetic cerebral palsy during powered mobility

**DOI:** 10.1101/2025.08.15.25333752

**Authors:** Saranda Bekteshi, Ioana Gabriela Nica, Benoit Cuyvers, Sotirios Gakopoulos, Hans Hallez, Elegast Monbaliu, Jean-Marie Aerts

## Abstract

**Background:** Dyskinetic cerebral palsy (DCP) is dominated by dystonia and choreoathetosis, two movement disorders that are often simultaneously present and challenging to evaluate. Wearable technology shows potential for monitoring motor dysfunctions at high temporal resolution while expanding our understanding of DCP movement disorders.

**Objectives:** This study aimed (i) to develop a methodology for automatic classification of dystonia and choreoathetosis combining inertial measurement units (IMUs) and random forests (RFs) during powered wheelchair driving in participants with DCP, (ii) to determine signature features for dystonia and choreoathetosis, and (iii) to optimise placement of body-worn IMUs in function of dystonia and choreoathetosis classification performance.

**Methods:** Unconstrained movements of the arms and head during powered mobility (n = 5 DCP participants) were analysed to extract 111 time- and frequency-domain features in 5-second windows. RFs were then used to rank, select optimal features and classify dystonia and choreoathetosis, based on expert-annotated videos.

**Results:** Classification of dystonia and choreoathetosis for the neck, proximal and distal arm regions ranged within 67.8% - 80.7% accuracy. Reduced feature sets included between 19 - 73 features, as time-domain features were selected more prevalently in classifying both dystonia and choreoathetosis. IMUs on the distal arms predicted forehead dystonia and choreoathetosis with similar accuracy (74.5% - 81.2%) as using the forehead IMU.

**Conclusions:** This study increases insights into DCP by relating distinct IMU features to dystonia and choreoathetosis and by leveraging distal arm-placed IMUs to assess movement disorders in multiple body parts: distal arm, proximal arm and neck region.

## Background

The development of smart wearables that quantify motor dysfunction can help expand current clinical assessments to allow automated monitoring of patients’ status in various contexts and environments. Efforts to develop such devices and algorithms are still in the inception phase for cerebral palsy (CP). CP is the most prevalent source of severe motor impairment in childhood^1^. Dyskinetic cerebral palsy (DCP) forms the second largest CP group after spastic CP^2^, and constitutes the most disabling subtype^3^. DCP is dominated by two movement disorders, dystonia and choreoathetosis, which are mostly simultaneously present and can vary in time and between body parts^2, 4^. Dystonia is more prominent and presents as twisting, repetitive movements and abnormal postures^2, 4, 5^. Choreoathetosis is a combination of chorea - rapid, jerky, often fragmented involuntary movements and athetosis - slower, continuous, involuntary writhing movements^2, 4, 5^. More than 70% of DCP patients present with the highest levels of severity in gross motor functioning and fine manual abilities^5^. As a result, the majority of children with DCP are non-ambulant and unable to steer powered wheelchairs with a conventional joystick^6^. Despite severe impairments, children with DCP can be trained to drive head/foot steering wheelchairs^7^. Still, the learning process can be very long and training with a wheelchair that does not meet the needs of the user may lead to frustration and abandonment. Since dystonia and choreoathetosis are the most disabling factors in the training process^7^, tracking their expression in real-time, while driving, is essential to adjust wheelchair settings to user needs and possibly improve the learning process.

Wearable sensors, such as inertial measurement units (IMUs), record subjects’ movements and can be used in combination with machine learning to detect presence and label severity of movement disorders. Such setups have been previously implemented to evaluate upper limb motor function in DCP^8^, stroke subjects^9^, to monitor freeze of gait in Parkinson’s disease (PD)^10^, to assess movement impairment in Huntington’s disease^11^ and to evaluate spasticity in children with spastic CP^12^. A few studies have attempted to track movement disorders during continuous, unconstrained activities with this technology, while focusing on tremor and dyskinesia in PD^13, 14^. Despite advances, no system is currently available to automatically monitor CP or specifically DCP movement disorders during daily activities like wheelchair driving. However, a rating protocol, the Dyskinesia Impairment Scale (DIS), was developed specifically for the DCP population to map the occurrence and severity of dystonia and choreoathetosis over different body regions^15^. The Dyskinesia Impairment Mobility Scale (DIMS), an adapted protocol of the DIS to quantify dystonia and choreoathetosis during powered mobility, was used as the reference standard for continuous labelling during head/foot steering in participants with DCP^16^.

No validated system currently provides automated, real-time monitoring of DCP movement disorders during powered mobility, i.e., the most relevant scenario for wheelchair training and interface personalisation. Prior work in head/foot steering has shown that the use of wearable IMUs during head/foot steering is feasible with participants with DCP; and that dystonia is closely related to exercise load, whereas choreoathetosis is associated with physical activity intensity and energy expenditure, underscoring the importance of monitoring both movement disorders during driving^17^. Moreover, the fluctuating nature of dystonia and choreoathetosis manifestation suggests value in cross-region sensing strategies (e.g., whether distal limb IMUs can inform neck dystonia during driving) to minimise instrumentation burden in real-world use.

Therefore, in this study, we describe a methodology for automatic classification of dystonia and choreoathetosis based on IMUs and random forests (RFs) during unconstrained powered mobility in individuals with DCP. We further investigate the feasibility of using distal arm-mounted IMUs to infer dyskinesia in the neck region, aiming to inform future strategies for more practical, low-burden monitoring systems.

## Methods

### Participants

The study group was previously described^7, 15, 17, 18^. Briefly, five participants aged 6-21 years old (three females) were recruited from two special education schools for children with motor disorders. Since we intended to investigate severe dystonia and choreoathetosis in CP, we included children diagnosed with DCP and categorised as levels IV-V for both gross motor abilities classified with the Gross Motor Function Classification System Expanded&Revised (GMFCS E&R)^19^ and fine manual abilities classified with the Manual Abilities Classification System (MACS)^20^. Only children who were users of a head/foot steering wheelchair were selected. Exclusion criteria included: difficulty in understanding and following instructions, severe visual impairment or a history of orthopaedic or neurosurgical interventions within the last 12 months. The study was approved by the Medical Ethics Committee UZ KU Leuven and conducted in accordance with the Declaration of Helsinki. Informed signed consent and assent were acquired from all participants and their primary caregivers.

### Data acquisition

We recorded data during five representative powered mobility tasks: independent driving, driving through a created corridor, a 360-degree turn to the left, a 360-degree turn to the right and a slalom as previously described^15^. Each participant performed the tasks twice, two weeks apart. All tests were carried out in the schools of the five participants using their personal head/foot steering wheelchair. The tests were video-recorded with two Sony Handycam HDR-CX405 (Sony Corporation, Tokyo, sampling frequency of 29 frames per second), placed on a tripod on both sides of the powered mobility setup.

Our goal was to quantify involuntary movements of the participants while operating a moving wheelchair. For this purpose, we attached five Shimmer3 IMUs (Shimmer Sensing, Dublin, Ireland) with 3D accelerometer and gyroscope to the participants’ distal arms, shins and forehead. Additionally, one IMU was placed at the back of the head support set as the master IMU, to automatically synchronise the other IMUs to its clock. The data logging method developed for this setup was previously published^18^. Data was recorded at a sampling frequency of 51.28 Hz, logged on the SD cards of each IMU during the tests and later transferred to a laptop for offline processing. As supported by clinical practice, participants’ feet were strapped to the wheelchair supports to minimise the effect of legs’ involuntary movements during driving. Because we focused this study on unconstrained movements, we analysed only data from the forehead- and the distal arm-attached IMUs.

### Data analysis

Based on DIMS scores and IMU data, we trained random forests (RFs) to classify levels of dystonia and choreoathetosis, but also to rank feature importance. All IMU analyses were conducted offline in MATLAB 2019b (MathWorks, Inc., Natick, MA, USA). The overall analysis and the classification scenarios are detailed in Figure 1.A.

**Figure 1.**
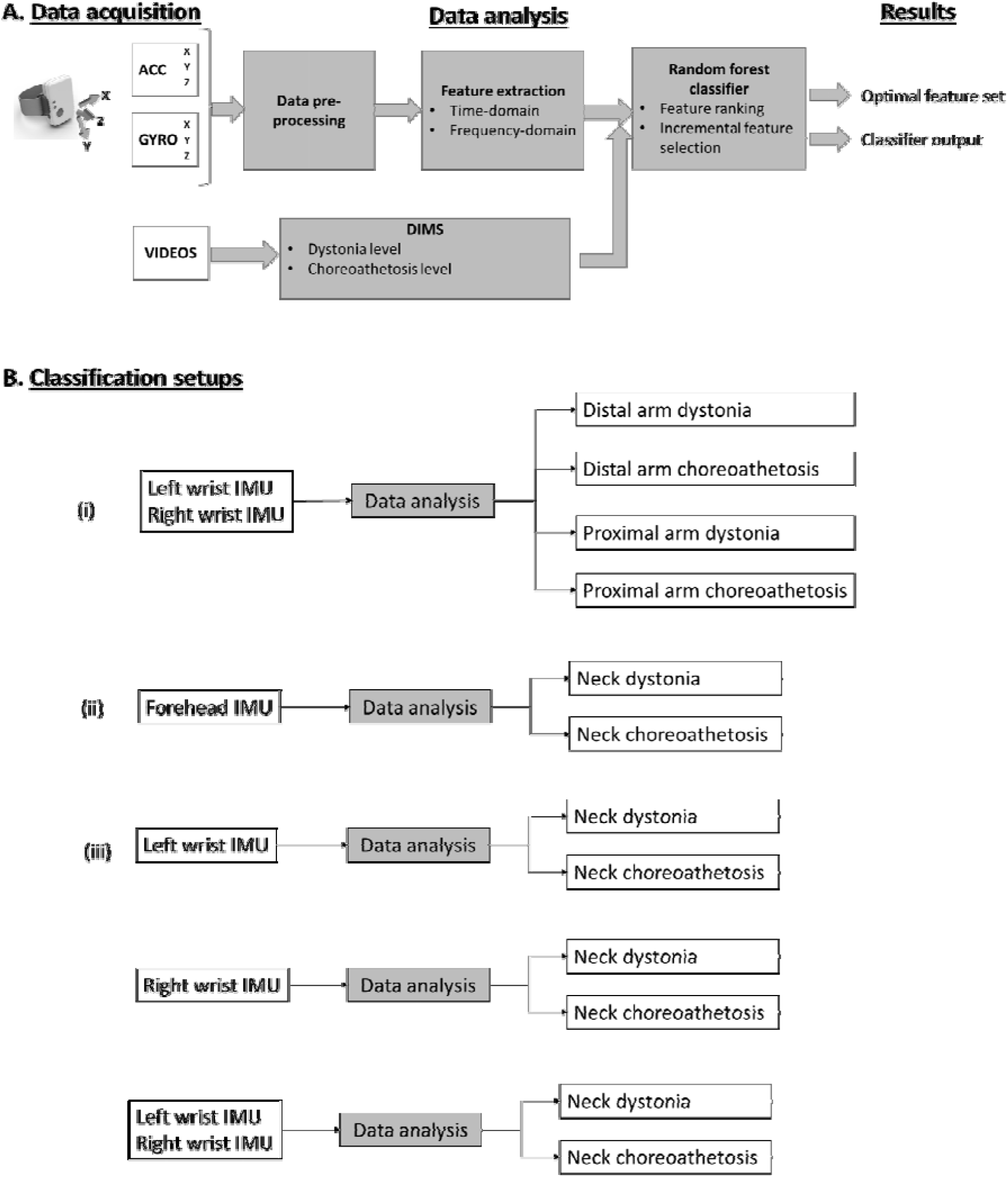
**A**. Overview of data analysis steps to classify levels of dystonia and choreoathetosis and determine 390 optimal feature sets for the classification. **B**. Classification setups used in this paper.

#### Video labels

The video recordings of each participant were manually scored according to the DIMS^14^. An experienced rater scored driving recordings over 5-second windows. Dystonia and choreoathetosis were scored separately for the proximal and distal parts of the upper limbs as well as the neck. The DIMS instrument uses a 5-point scale for amplitude, between 0 = absence of the movement disorder, to 4 = movement disorder is present in the maximal range of motion.

#### IMU pre-processing

During the experiments, IMUs were turned on and off manually. Each sensor established a connection with the master IMU and started transmitting individually. The communication protocol synchronised the sensors automatically. However, to ensure perfect timeline overlap, the small differences in the start time of each sensor were removed. Next, we manually synchronised the IMU recordings with the video footage. While recording, small data losses occurred due to weak signal transmission. The maximum loss was 7 consecutive samples (∼136 ms), and in total, less than 0.4% of the recordings were lost. The signal was reconstructed using linear interpolation. After subtraction, IMU data was low-pass filtered using a Butterworth filter (cutoff frequency 17 Hz, optimal order determined by Matlab function “buttord”) and downsampled to 13.89 Hz. The cutoff frequency was chosen based on existing reports that voluntary and involuntary movements are localised within the bandwidth 0-4 Hz^21, 22^. Finally, exercise intervals were extracted and divided into 5-second windows.

#### IMU features

##### Time-domain features

Abnormal postures are often reported in dystonia^2^. Postures can be encoded, especially in the time-domain of accelerometer data. We included root mean square (RMS), variance, as well as pairwise correlations between the three-dimensional acceleration and gyroscope signals, using the Pearson correlation coefficient. Accelerometer autocorrelation has been used to identify the dominant frequency of tremor in PD^13^. Choreoathetosis is associated with jerky involuntary movements, so in order to quantify (lack of) movement smoothness, we extracted RMS jerk and dimensionless jerk^23^. Given the characteristics of dystonia and choreoathetosis, we also included complexity measures: permutation entropy^24^, to measure the absence of permutation patterns within a time series and sample entropy^25^, to assess the complexity and regularity within the time series data, as it measures the degree of dependency of a given data point on a number of previous data points.

##### Frequency-domain features

Since dystonia manifests as twisting, repetitive movements, we hypothesised that the frequency-domain of both accelerometer and gyroscope data encoded the rhythmicity of the twisting movements. Choreoathetosis movements have been reported to present higher frequencies (>1 Hz^22^, 2 – 4 Hz^21^) than voluntary movements (0.3-0.5 Hz)^22^, but in general, involuntary and voluntary movements have the same bandwidth (0-4 Hz)^19^.We hypothesised that chorea could be detected as sharp, higher frequency oscillations in accelerometer and gyroscope data. The frequency-domain features were extracted using the continuous wavelet transform on accelerometer and gyroscope data, with the Morlet family as its basis. In each 5-second window, we identified instantaneous frequency peaks, as well as the mean value, the variance and the maximum value of the strongest frequencies. To quantify the dynamics of the frequency spectrum, we also applied the discrete wavelet transform (DWT) to the accelerometer and gyroscope data. The resulting detail and approximation coefficients cD1 (3.5-7 Hz), cD2 (1.75 – 3.5 Hz), cD3 (0.85-1.75 Hz) and cA3 (0-0.85 Hz) are a measure of the energy in their frequency band. We calculated the squared coefficients for each frequency band, and we extracted the percentage of energy for each frequency band by dividing its energy level by the sum of all energy bands. Finally, we also included entropy measures to assess the degree of chaos in the frequency-domain of these signals using spectral entropy^26^. Finally, exercise intervals were extracted and divided into 5-second windows. Table 1 provides an overview of the time- and frequency-domain features extracted.

**Table 1.** Summary of time-domain and frequency-domain features.

| Feature | No. of attributes per window<sup>a</sup> | Feature description | Hypothesized related symptom<sup>b</sup> |
| --- | --- | --- | --- |
| RMS (RMS_ag) | 6 | 1D movement and postural control | D & CA |
| Variance (var_ag) | 6 | Variability | D & CA |
| RMS jerk (RMS_jerk_ag) | 6 | Smoothness of movement | CA |
| Dimensionless jerk (dim_jerk_ag) | 6 | Smoothness of movement | CA |
| Variance jerk (var_jerk_ag) | 6 | Variability | CA |
| Pairwise correlation between axes (ag_xyz) | 15 | 2D movements and postural control | D & CA |
| Permutation entropy (PE_ag) | 6 | Complexity in time | CA |
| Sample entropy (SaE_ag) | 6 | Complexity in time | CA |
| Spectral entropy (SpE_ag) | 6 | Complexity in frequencies | CA |
| Number of peaks (numpeaks_ag) | 6 | Smoothness of movement | D |
| Mean of strongest frequencies (mean_freq_ag) | 6 | Specific rhythmicity | D |
| Variance of strongest frequencies (var_freq_ag) | 6 | Smoothness of movement | CA |
| Maximum of strongest frequencies (max_freq_ag) | 6 | Specific rhythmicity | D |
| Energy band D1 coefficient (fbandD1_ag) | 6 | Energy between 3.5 – 7 Hz | CA |
| Energy band D2 coefficient (fbandD2_ag) | 6 | Energy between 1.7 – 3.5 Hz | CA |
| Energy band D3 coefficient (fbandD3_ag) | 6 | Energy between 0.8 – 1.7 Hz | D & CA |
| Energy band A3 coefficient (fbandA3_ag) | 6 | Energy between 0 – 0.8 Hz | D |

<sup>a</sup>The number of attributes per time window generally corresponds to one value per axis, per time window (i.e. ‘ag’ signifies one attribute for each of the 3 axes of the acceleration and, respectively, the angular velocity).
<sup>b</sup>Hypothesized decoding relevance in relation to dystonia (D) and/or choreoathetosis (CA).

#### Random forest classification and optimal feature selection

The feature extraction resulted in 111 time- and frequency-domain feature values per 5-second window. In the next step, we used RFs to gain insight into feature importance when classifying levels of dystonia and choreoathetosis. RFs constitute a popular machine-learning technique for approaching different biological prediction problems, like IMU-based movement analysis^27-29^, gene-technology^30, 31^, EEG signal processing^32^. Developed by Leo Breiman^33^, RF is an ensemble predictor that consists of multiple decision trees. Each tree is built using a bootstrap sample of the data. To classify a new queried sample, each tree provides a predicted class, and the outcome is the class with the most votes. RFs consider the importance of each feature individually and the interactions with other features, so that no pre-selection of features is needed. This technique is robust to overfitting and suitable for large variable sets when few observations are available. Random forests provide a measure of error rate based on the out-of-bag (OOB) cases for each fitted tree, the OOB error, and in this study, we selected the optimal feature set as the set minimising the OOB error rate.

We trained a different RF for each of the 12 classification scenarios described in Figure 1.B., so that dystonia and choreoathetosis were classified independently of each other in the three body regions of interest, neck, distal arms and proximal arms. We used RFs with an ensemble of 200 trees and permuting out-of-bag observations to rank the 111 features from higher to lower importance. Next, ranked features were added one by one from higher to lower rank to determine the optimal number of features^28^. A new feature set was composed when one feature was added, and for each of the 111 sets of features, an RF was constructed and the OOB error was estimated. We selected the feature subset that achieved the best prediction performance. Finally, we calculated the prediction sensitivity, specificity and accuracy for the optimal, reduced feature set.

## Results

### DIMS scores distribution and classification labels

Video recordings of driving tasks were analysed by an experienced rater who evaluated 303 five-second epochs. Separate amplitude scores were provided for dystonia and choreoathetosis in the neck, proximal arms and distal arms according to the DIMS protocol. In case the body part was not clearly visible or the resolution of the image did not allow for an evaluation, the epoch was eliminated. Overall, it was possible to score 80.5% of the neck data, 61.9% of the distal arm data and 79.5% of the proximal arm data.

The distribution of the five amplitude scores was uneven (Figure 2), with predominantly high amplitude scores in dystonia (86.4%, 64.4%, 21.9%) and a high proportion of scores 0 in choreoathetosis (50.1%, 53.1%, 31.4%). To create more balanced datasets suitable for classification, we regrouped the samples into new labels (modified DIMS score, Figure 2). For choreoathetosis, this resulted in a two-class distribution, where 0 indicates the absence of choreoathetosis and 1 indicates the presence of choreoathetosis (DIMS levels 1-4). For dystonia, we created a three-class distribution, where 0 indicates absence of dystonia (DIMS score 0), 1 indicates low presence of dystonia (DIMS scores 1-3 for arms and DIMS score 1 for neck) and 2 indicates strong presence of dystonia (DIMS score 4 for arms and DIMS scores 2-4 for neck).

**Figure 2.**
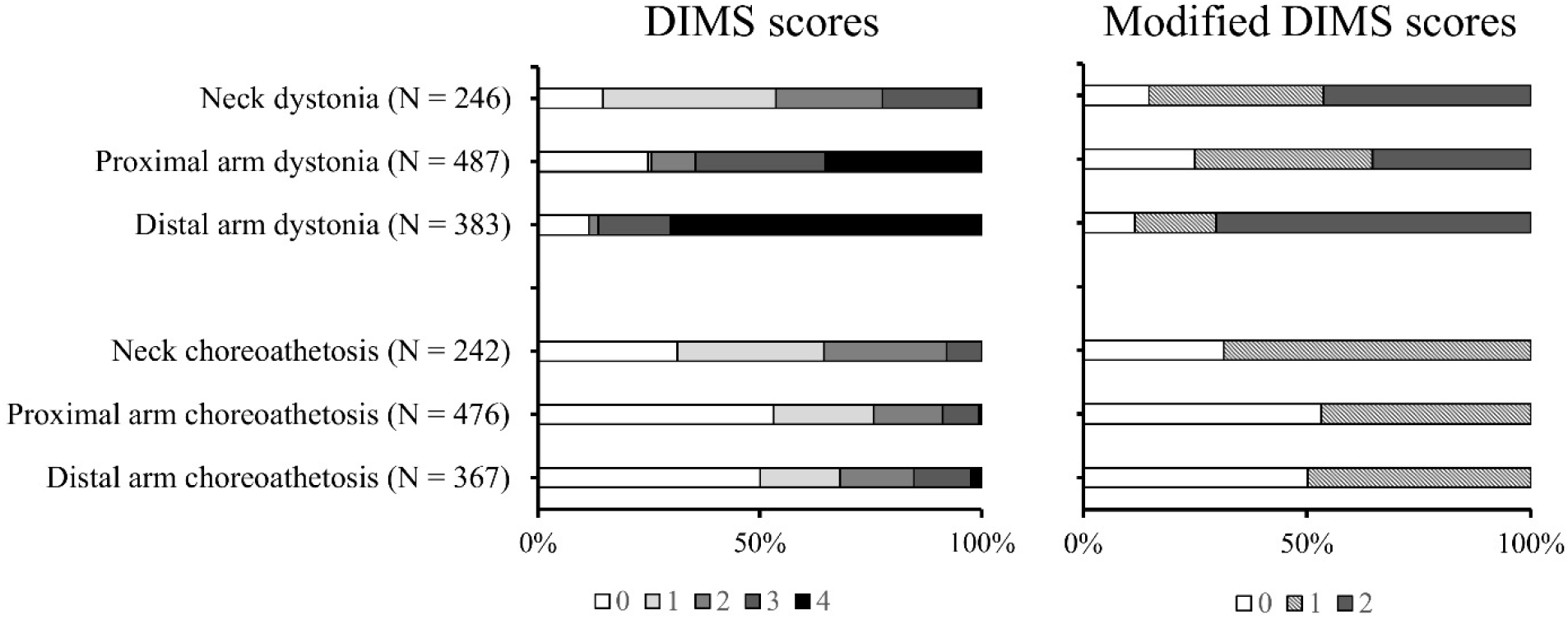
Dyskinesia prevalence (%) based on DIMS scores (left) and modified dyskinesia scores (right).

### Classification of dystonia and choreoathetosis

The modified DIMS scores were the gold standard for the classification of dystonia and choreoathetosis, per body region. In each classification scenario described in Figure 1, we created 111 RF classifiers for all sub-feature sets and we selected the sets of features generating minimum OOB error. The complete overview of the optimal feature sets is provided in Supplementary Table 1.

Signal features from the right and left distal arm IMUs were fused for classifying distal arm dystonia (prediction accuracy 78.7% with 16 features), distal arm choreoathetosis (accuracy 67.8% with 54 features), proximal arm dystonia (accuracy 73.9% with 55 features) and proximal arm choreoathetosis (accuracy 75.8% with 73 features), respectively. IMU features from the forehead sensors were used for classifying neck dystonia, achieving a prediction accuracy of 80.1% with 43 features and neck choreoathetosis, with an accuracy of 76.3% based on 19 features.

### Analysis of optimal feature sets

We grouped the selected feature sets based on two criteria: (i) the sensor data originated from an accelerometer versus gyroscope data, and (ii) the variable with respect to which data was analysed into time-versus frequency-domain features. Their distributions are shown in Figure 3, with no distinction made between body regions. Slightly more accelerometer features were selected for the dystonia classifiers (56.1% of total features), while more gyroscope features were retained for the choreoathetosis classification (53.4% of total features), but overall data from both sensors proved important in the classification. Time-domain features were dominant in classifying both movement disorders, with 66/114 features for dystonia and 87/146 features selected for the choreoathetosis classifiers. Accelerometer RMS was more prevalent in dystonia quantification (66.7% occurrence) than in choreoathetosis classification (38.9% occurrence). Variance (50% occurrence in dystonia and 55.6% in choreoathetosis) and jerk-related features (55.6% occurrence in dystonia and 70.4% in choreoathetosis) were frequently selected in the classification of both disorders. From the set of complexity measures, permutation entropy (33.3% occurrence) was most selected for dystonia classification, while sample entropy (61.1%) and spectral entropy (44.4%) were more often selected for choreoathetosis. Frequency peaks and dynamics of the power spectra were selected more often for choreoathetosis (33.3 % versus 44.4% and 16.7% versus 35.2%). The distribution of energy per frequency bands was comparable for both disorders.

**Figure 3.**
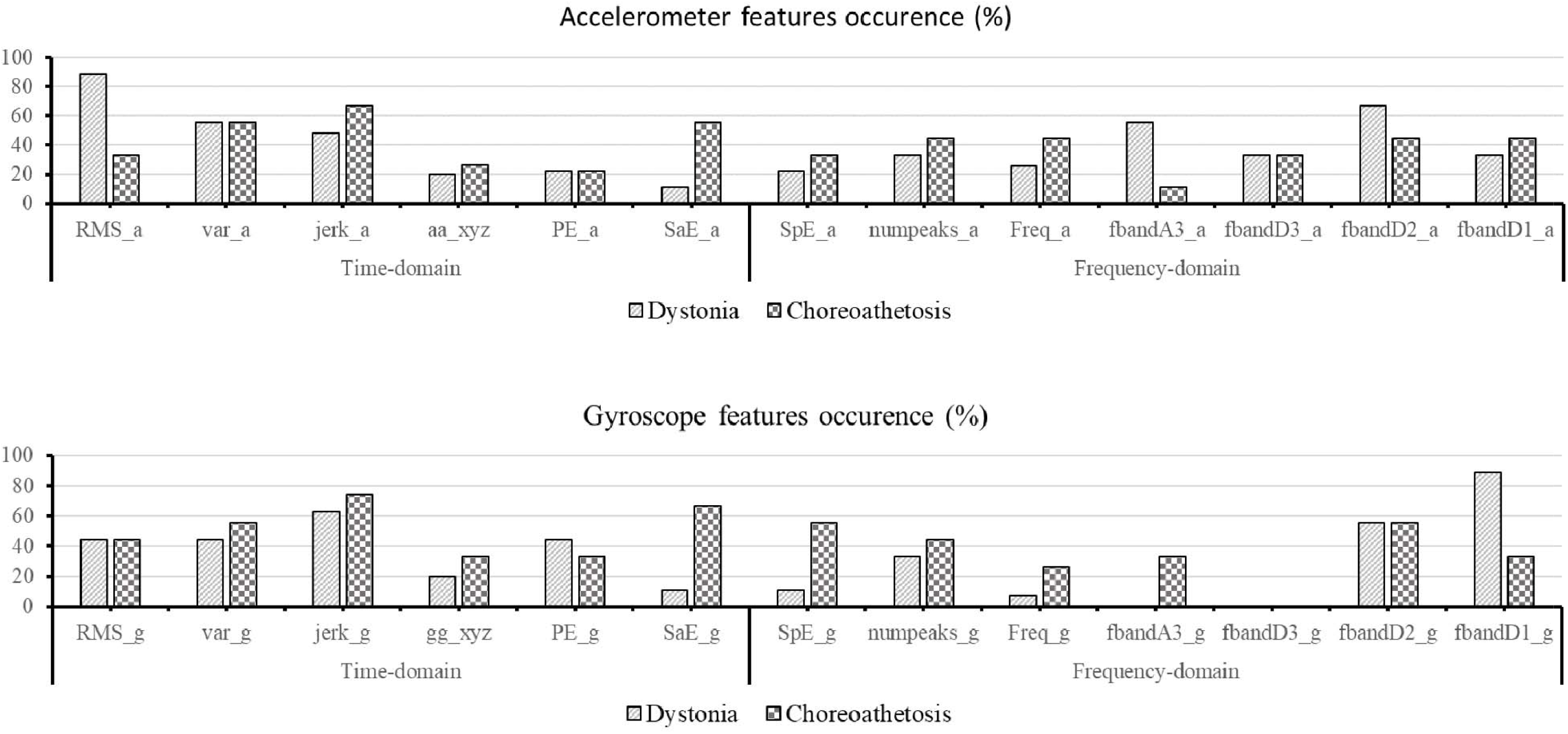
Comparison of accelerometer (top) and gyroscope (bottom) features.

### Distal arm IMUs for neck choreoathetosis and dystonia

As described, signal features of the forehead IMU were used to classify dystonia and choreoathetosis in the neck region. However, in DCP, dystonia and choreoathetosis have been shown to manifest as overflow movements (unwanted movement associated with an intended movement) at significantly higher levels in extremities than in the central body^34^. Thus, we investigated the potential relation between involuntary movements in the neck and distal arms regions by training classifiers for neck dystonia and choreoathetosis using signal features of the arms IMUs. The performance of the selected feature sets was evaluated by comparing the sensitivity, specificity, and accuracy of the classifiers, as shown in Table 2. Individual arm IMUs or a combination of both datasets could be used to generate comparable results to using the forehead IMU data. However, in general, better results were achieved with data from one distal arm rather than a combination of both. In particular, the neck choreoathetosis classification resulted in high sensitivity, low specificity.

**Table 2.**
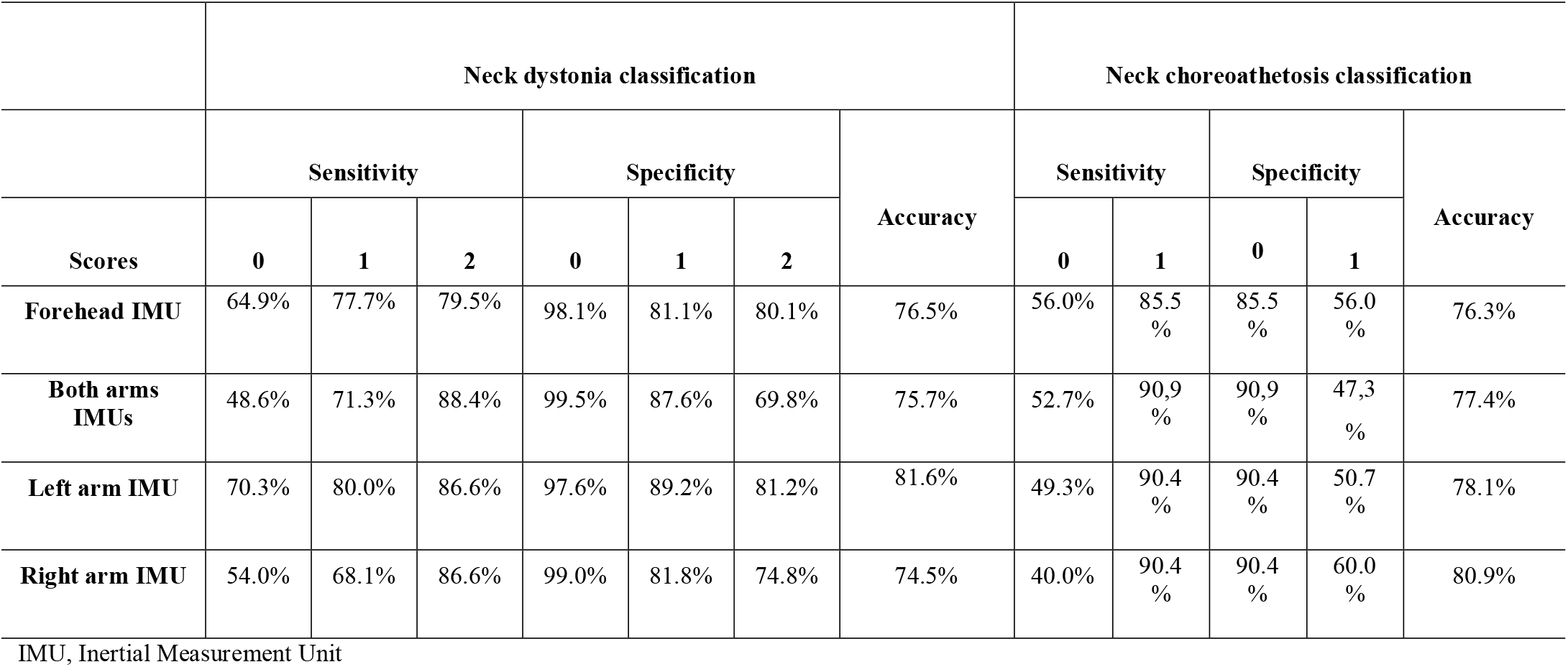
Comparison of classifiers performance using either forehead IMU or arms IMU data for dystonia and choreoathetosis classification.

## Discussion

This paper is one of the earliest that focused on quantifying dystonia and choreoathetosis with wearable sensors in CP participants. A unique value is the use of wearable sensors during powered mobility with head/foot steering, a functionally meaningful yet technically challenging context. We developed a methodology to identify the optimal signal features to classify dystonia and choreoathetosis during powered mobility. The selection of relatively large feature sets reflected the complexity of detecting these movement disorders. Indeed, we achieved lower classification performance than existing studies on wearables that report sensitivity and specificity of 90% or better in detecting dyskinesia in PD, some under added constraints, like scripted-activity monitoring conditions^35-37^ or lower temporal resolution^36^. A few studies monitored continuous unconstrained activities, using one sensor per limb of interest^13, 14^. However, our approach is unique in that we showed that distal arm-attached IMUs can be leveraged to assess three body regions: the distal and proximal arm as well as the neck regions. The uneven distribution of the DIMS amplitude scores (Figure 2) created a bias in the classification, which decreased the generalisation potential of the classifiers. Since we evaluated motor limitations in wheelchair-bound DCP participants, we included in this study only children classified with levels IV-V for both GMFCS and MACS, which explains the high prevalence of severe dystonia.

Potentially useful features were extracted from accelerometer and gyroscope data, and their significance in classifying levels of dystonia and choreoathetosis was assessed. Both sensors contributed to the classification of the two movement disorders, with linear acceleration features slightly more prevalent in the analysis of dystonia and rotational features slightly more often selected in the analysis of choreoathetosis (Figure 3). Expansion of the data set is needed to confirm this discrepancy. Overall, time-domain features were more prevalent, suggesting they can be used to represent the movement patterns specific to dystonia chorea better than frequency-domain features, in agreement with a previous study in Huntington’s disease^11^. High prevalence of selecting accelerometer RMS (88.9 % occurrence) in dystonia classifiers suggests translational accelerations are more related to the manifestation of dystonia. High prevalence of variances and jerk variables from both sensors reflect that unsmooth translational and rotational movements are present in both dystonia and choreoathetosis. Our hypothesis regarding a possible discrepancy into lower frequency representation for dystonia and higher frequency representation for choreoathetosis was not confirmed. Despite existing literature, we found dystonia to be related to higher frequencies (1.7-3.5 Hz).

Participants steer the head-foot wheelchair system using their head. Thus, monitoring neck dystonia and neck choreoathetosis is crucial to supervise driving ability. At the same time, the forehead-attached IMU was reported during the tests as the most uncomfortable part of the experimental setup. In DCP, dystonia and choreoathetosis can also present as overflow movements induced in different body parts than those involved in goal-directed movements^34^. If this phenomenon occurs during wheelchair steering, involuntary movements in the arms could be overflow movements induced by pressing movements of the head. We classified neck dystonia and neck choreoathetosis using IMU signals recorded on the arms and with similar accuracy as using the forehead IMUs, as illustrated in Table 2. This opens the potential of using wrist-worn sensors to monitor involuntary movements of the head in this population. 34.5% (Supplementary Table 2) of dystonia features from the forehead IMU set were present in all three classification scenarios involving arms IMU data. Moreover, 93% of original dystonia features were present in at least one of the three distal arm feature sets, suggesting similar mechanisms of dystonia manifestation between body parts. This was strikingly not the case with choreoathetosis, where only 5% of forehead IMU forehead features were selected in all three classification scenarios involving arms IMU data, while 31% of forehead IMU variables were never selected.

Since this study was conducted, further work has expanded the field: reliability and discriminative validity of IMU-based features in DCP have been established^8^, home-based smartphone-coupled IMU monitoring has demonstrated feasibility^38^, video-based automated approaches have shown promise^39^, particularly for dystonia detection, and the DIS-II has streamlined clinical rating^40^. These developments contextualise our work as an important step in the transition from feasibility to broader clinical implementation, especially for powered mobility, where real-time monitoring could inform adaptive wheelchair control strategies.

Future research should extend to larger and more diverse cohorts, cover the full range of DIMS severity levels, and integrate multi-modal data sources (such as IMUs, video, and physiological signals) to improve accuracy. Ultimately, embedding such monitoring into adaptive assistive technologies could personalise training, enhance user engagement, reduce technology discontinuance, and foster meaningful gains in functional activities, participation, and overall quality of life.

## Conclusions

The approach presented in this study demonstrated for the first time the use of IMUs for the detection of dystonia and choreoathetosis in CP participants. We identified optimal signal features to classify these two movement disorders in the distal and proximal arms and neck regions, while the participants were engaged in driving their wheelchairs. Our results indicate that time-domain features play a more prevalent role than frequency-based features in the classification of both disorders. Arm-attached IMUs could detect dystonia and choreoathetosis in the neck region with promising accuracy. Given these encouraging results, we are currently applying this methodology in characterising voluntary movements to describe driving patterns in the same patient group. Future research should focus on improving classification accuracy in an extended patient group.

## Data Availability

All data produced in the present study are available upon reasonable request to the authors.

## Availability of data and materials

Data will be made available upon request to the corresponding author.

## Acknowledgment

The authors express sincere gratitude to the participants involved in this study and to their supportive parents and therapists for their crucial help during the data collection.

---

**Supplementary Table 1.**
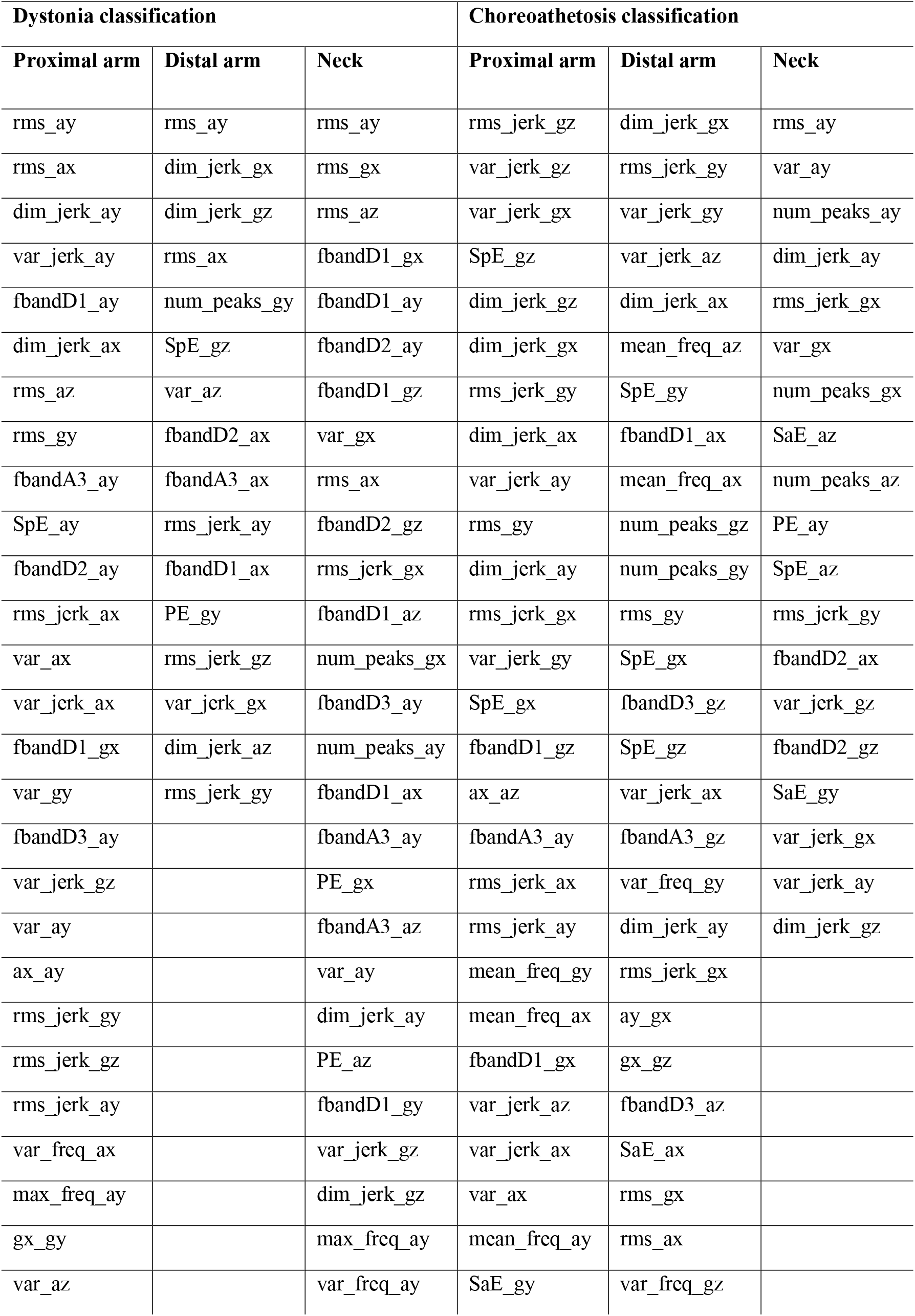
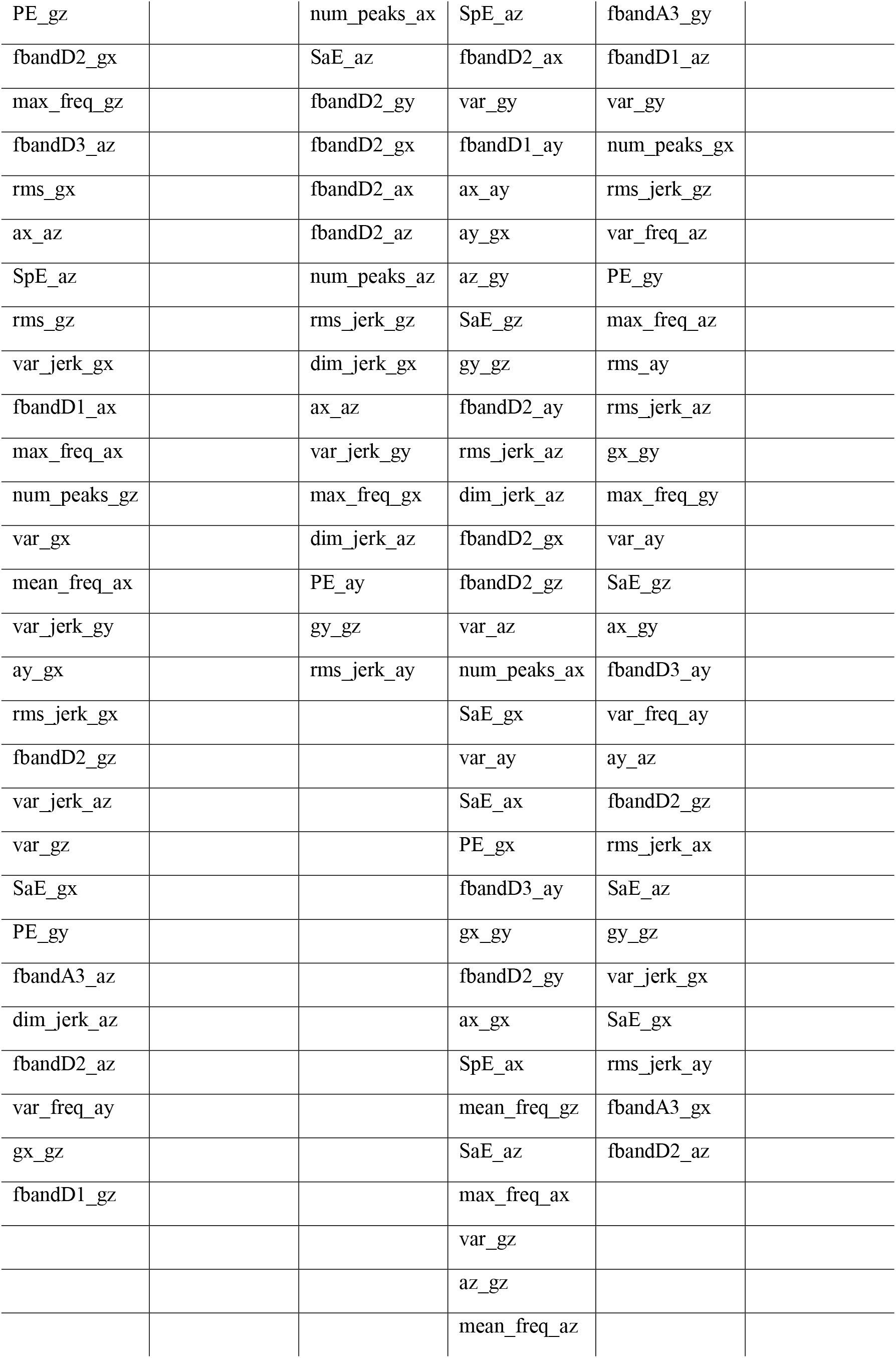
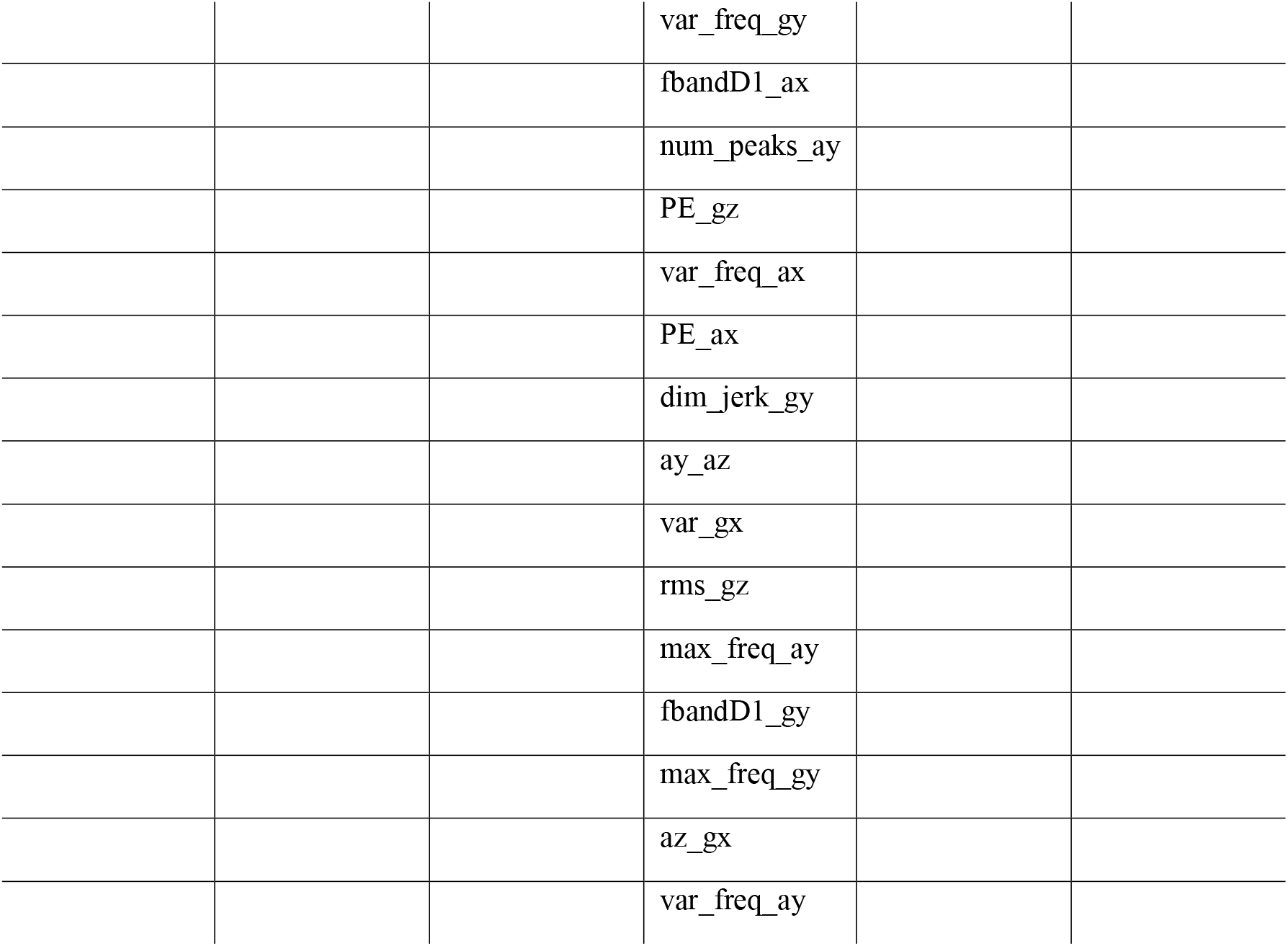
Ranking of relevant feature sets.

| **Dystonia classification** |  |  | **Choreoathetosis classification** |  |  |
| --- | --- | --- | --- | --- | --- |
| **Proximal arm** | **Distal arm** | **Neck** | **Proximal arm** | **Distal arm** | **Neck** |
| rms_ay | rms_ay | rms_ay | rms_jerk_gz | dim_jerk_gx | rms_ay |
| rms_ax | dim_jerk_gx | rms_gx | var_jerk_gz | rms_jerk_gy | var_ay |
| dim_jerk_ay | dim_jerk_gz | rms_az | var_jerk_gx | var_jerk_gy | num_peaks_ay |
| var_jerk_ay | rms_ax | fbandD1_gx | SpE_gz | var_jerk_az | dim_jerk_ay |
| fbandD1_ay | num_peaks_gy | fbandD1_ay | dim_jerk_gz | dim_jerk_ax | rms_jerk_gx |
| dim_jerk_ax | SpE_gz | fbandD2_ay | dim_jerk_gx | mean_freq_az | var_gx |
| rms_az | var_az | fbandD1_gz | rms_jerk_gy | SpE_gy | num_peaks_gx |
| rms_gy | fbandD2_ax | var_gx | dim_jerk_ax | fbandD1_ax | SaE_az |
| fbandA3_ay | fbandA3_ax | rms_ax | var_jerk_ay | mean_freq_ax | num_peaks_az |
| SpE_ay | rms_jerk_ay | fbandD2_gz | rms_gy | num_peaks_gz | PE_ay |
| fbandD2_ay | fbandD1_ax | rms_jerk_gx | dim_jerk_ay | num_peaks_gy | SpE_az |
| rms_jerk_ax | PE_gy | fbandD1_az | rms_jerk_gx | rms_gy | rms_jerk_gy |
| var_ax | rms_jerk_gz | num_peaks_gx | var_jerk_gy | SpE_gx | fbandD2_ax |
| var_jerk_ax | var_jerk_gx | fbandD3_ay | SpE_gx | fbandD3_gz | var_jerk_gz |
| fbandD1_gx | dim_jerk_az | num_peaks_ay | fbandD1_gz | SpE_gz | fbandD2_gz |
| var_gy | rms_jerk_gy | fbandD1_ax | ax_az | var_jerk_ax | SaE_gy |
| fbandD3_ay |  | fbandA3_ay | fbandA3_ay | fbandA3_gz | var_jerk_gx |
| var_jerk_gz |  | PE_gx | rms_jerk_ax | var_freq_gy | var_jerk_ay |
| var_ay |  | fbandA3_az | rms_jerk_ay | dim_jerk_ay | dim_jerk_gz |
| ax_ay |  | var_ay | mean_freq_gy | rms_jerk_gx |  |
| rms_jerk_gy |  | dim_jerk_ay | mean_freq_ax | ay_gx |  |
| rms_jerk_gz |  | PE_az | fbandD1_gx | gx_gz |  |
| rms_jerk_ay |  | fbandD1_gy | var_jerk_az | fbandD3_az |  |
| var_freq_ax |  | var_jerk_gz | var_jerk_ax | SaE_ax |  |
| max_freq_ay |  | dim_jerk_gz | var_ax | rms_gx |  |
| gx_gy |  | max_freq_ay | mean_freq_ay | rms_ax |  |
| var_az |  | var_freq_ay | SaE_gy | var_freq_gz |  |
| PE_gz |  | num_peaks_ax | SpE_az | fbandA3_gy |  |
| fbandD2_gx |  | SaE_az | fbandD2_ax | fbandD1_az |  |
| max_freq_gz |  | fbandD2_gy | var_gy | var_gy |  |
| fbandD3_az |  | fbandD2_gx | fbandD1_ay | num_peaks_gx |  |
| rms_gx |  | fbandD2_ax | ax_ay | rms_jerk_gz |  |
| ax_az |  | fbandD2_az | ay_gx | var_freq_az |  |
| SpE_az |  | num_peaks_az | az_gy | PE_gy |  |
| rms_gz |  | rms_jerk_gz | SaE_gz | max_freq_az |  |
| var_jerk_gx |  | dim_jerk_gx | gy_gz | rms_ay |  |
| fbandD1_ax |  | ax_az | fbandD2_ay | rms_jerk_az |  |
| max_freq_ax |  | var_jerk_gy | rms_jerk_az | gx_gy |  |
| num_peaks_gz |  | max_freq_gx | dim_jerk_az | max_freq_gy |  |
| var_gx |  | dim_jerk_az | fbandD2_gx | var_ay |  |
| mean_freq_ax |  | PE_ay | fbandD2_gz | SaE_gz |  |
| var_jerk_gy |  | gy_gz | var_az | ax_gy |  |
| ay_gx |  | rms_jerk_ay | num_peaks_ax | fbandD3_ay |  |
| rms_jerk_gx |  |  | SaE_gx | var_freq_ay |  |
| fbandD2_gz |  |  | var_ay | ay_az |  |
| var_jerk_az |  |  | SaE_ax | fbandD2_gz |  |
| var_gz |  |  | PE_gx | rms_jerk_ax |  |
| SaE_gx |  |  | fbandD3_ay | SaE_az |  |
| PE_gy |  |  | gx_gy | gy_gz |  |
| fbandA3_az |  |  | fbandD2_gy | var_jerk_gx |  |
| dim_jerk_az |  |  | ax_gx | SaE_gx |  |
| fbandD2_az |  |  | SpE_ax | rms_jerk_ay |  |
| var_freq_ay |  |  | mean_freq_gz | fbandA3_gx |  |
| gx_gz |  |  | SaE_az | fbandD2_az |  |
| fbandD1_gz |  |  | max_freq_ax |  |  |
|  |  |  | var_gz |  |  |
|  |  |  | az_gz |  |  |
|  |  |  | mean_freq_az |  |  |
|  |  |  | var_freq_gy |  |  |
|  |  |  | fbandD1_ax |  |  |
|  |  |  | num_peaks_ay |  |  |
|  |  |  | PE_gz |  |  |
|  |  |  | var_freq_ax |  |  |
|  |  |  | PE_ax |  |  |
|  |  |  | dim_jerk_gy |  |  |
|  |  |  | ay_az |  |  |
|  |  |  | var_gx |  |  |
|  |  |  | rms_gz |  |  |
|  |  |  | max_freq_ay |  |  |
|  |  |  | fbandD1_gy |  |  |
|  |  |  | max_freq_gy |  |  |
|  |  |  | az_gx |  |  |
|  |  |  | var_freq_ay |  |  |

**Supplementary Table 2.** Comparison of feature sets.

|  | Head IMU for ND | Arms IMUs for ND | LA IMU for ND | RA IMU for ND |
| --- | --- | --- | --- | --- |
| # Features | 43 | 49 | 34 | 93 |
| Retained features (%) | - | 49.0 % | 52.9 % | 41.9 % |
|  | Head IMU for NC | Arms IMUs for NC | LA IMU for NC | RA IMU for NC |
| # Features | 19 | 35 | 8 | 34 |
| Retained features (%) | - | 25.7 % | 25.0 % | 20.6 % |

ND = neck dystonia; NC = neck choreoathetosis; LA = left arm; RA = right arm.

## References

1. Bekteshi S, Monbaliu E, McIntyre S, Saloojee G, Hilberink SR, Tatishvili N, et al. Towards functional improvement of motor disorders associated with cerebral palsy. The Lancet Neurology. 2023;22(3):229–43.

2. Monbaliu E, Himmelmann K, Lin JP, Ortibus E, Bonouvrie L, Feys H, et al. Clinical presentation and management of dyskinetic cerebral palsy. Lancet Neurol. 2017;16(9):741–9.

3. Préel M, Rackauskaite G, Larsen ML, Laursen B, Lorentzen J, Born AP, et al. Children with dyskinetic cerebral palsy are severely affected as compared to bilateral spastic cerebral palsy. Acta Paediatr. 2019;108(10):1850–6.

4. Monbaliu E, de Cock P, Ortibus E, Heyrman L, Klingels K, Feys H. Clinical patterns of dystonia and choreoathetosis in participants with dyskinetic cerebral palsy. Dev Med Child Neurol. 2016;58(2):138–44.

5. Monbaliu E, De La Pena MG, Ortibus E, Molenaers G, Deklerck J, Feys H. Functional outcomes in children and young people with dyskinetic cerebral palsy. Dev Med Child Neurol. 2017;59(6):634–40.

6. Arner M, Eliasson AC, Nicklasson S, Sommerstein K, Hägglund G. Hand function in cerebral palsy. Report of 367 children in a population-based longitudinal health care program. J Hand Surg Am. 2008;33(8):1337–47.

7. Bekteshi S, Konings M, Nica IG, Gakopoulos S, Aerts J-M, Hallez H, et al. Dystonia and choreoathetosis presence and severity in relation to powered wheelchair mobility performance in children and youth with dyskinetic cerebral palsy. European Journal of Paediatric Neurology. 2020.

8. Vanmechelen I, Bekteshi S, Haberfehlner H, Feys H, Desloovere K, Aerts J-M, et al. Reliability and Discriminative Validity of Wearable Sensors for the Quantification of Upper Limb Movement Disorders in Individuals with Dyskinetic Cerebral Palsy. Sensors. 2023;23(3):1574.

9. Li Y, Zhang X, Gong Y, Cheng Y, Gao X, Chen X. Motor Function Evaluation of Hemiplegic Upper-Extremities Using Data Fusion from Wearable Inertial and Surface EMG Sensors. Sensors (Basel). 2017;17(3).

10. Horak FB, Mancini M. Objective biomarkers of balance and gait for Parkinson’s disease using body-worn sensors. Movement disorders : official journal of the Movement Disorder Society. 2013;28(11):1544–51.

11. Bennasar M, Hicks YA, Clinch SP. Automated Assessment of Movement Impairment in Huntington’s Disease. IEEE Transactions on Neural Systems and Rehabilitation Engineering. 2018;26(10).

12. Choi S, Shin YB, Kim SY, Kim J. A novel sensor-based assessment of lower limb spasticity in children with cerebral palsy. J Neuroeng Rehabil. 2018;15(1):45.

13. Cole BT, Roy SH, De Luca CJ, Nawab SH. Dynamical learning and tracking of tremor and dyskinesia from wearable sensors. IEEE Trans Neural Syst Rehabil Eng. 2014;22(5):982–91.

14. Roy SH, Cole BT, Gilmore LD, Thomas CA, Saint-Hilaire M, Mayo C, et al. High-resolution tracking of motor disorders in Parkinson’s disease during unconstrained activity. Movement Disorders. 2013;28(8):1080–7.

15. Bekteshi S, Konings M, Nica IG, Gakopoulos S, Vanmechelen I, Aerts J-M, et al. Development of the Dyskinesia Impairment Mobility Scale to Measure Presence and Severity of Dystonia and Choreoathetosis during Powered Mobility in Dyskinetic Cerebral Palsy. Applied Sciences. 2019;9(17):3481.

16. Monbaliu E, Ortibus E, De Cat J, Dan B, Heyrman L, Prinzie P, et al. The Dyskinesia Impairment Scale: a new instrument to measure dystonia and choreoathetosis in dyskinetic cerebral palsy. Dev Med Child Neurol. 2012;54(3):278–83.

17. Bekteshi S, Nica IG, Gakopoulos S, Konings M, Maes R, Cuyvers B, et al. Exercise load and physical activity intensity in relation to dystonia and choreoathetosis during powered wheelchair mobility in children and youth with dyskinetic cerebral palsy. Disability and Rehabilitation. 2022;44(17):4794–805.

18. Gakopoulos S, Nica IG, Bekteshi S, Aerts JM, Monbaliu E, Hallez H. Development of a data logger for capturing human-machine interaction in wheelchair head-foot steering sensor system in dyskinetic cerebral palsy. Sensors (Switzerland). 2019;19(24).

19. Palisano RJ, Rosenbaum P, Bartlett D, Livingston MH. Content validity of the expanded and revised Gross Motor Function Classification System. Dev Med Child Neurol. 2008;50(10):744–50.

20. Eliasson AC, Krumlinde-Sundholm L, Rösblad B, Beckung E, Arner M, Ohrvall AM, et al. The Manual Ability Classification System (MACS) for children with cerebral palsy: scale development and evidence of validity and reliability. Dev Med Child Neurol. 2006;48(7):549–54.

21. Gresty MA, Halmagyi GM. Abnormal Head Movements. Journal of Neurology, Neurosurgery, and Psychiatry. 1979;42:705–14.

22. Raya R, Rocon E, Gallego JA, Ceres R, Pons JL. A Robust Kalman Algorithm to Facilitate Human-Computer Interaction for People with Cerebral Palsy, Using a New Interface Based on Inertial Sensors. Sensors (Basel). 2012;12(3):3049.

23. Hogan N, Sternad D. Sensitivity of Smoothness Measures to Movement Duration, Amplitude, and Arrests. 2010.

24. Bandt C, Pompe B. Permutation Entropy: A Natural Complexity Measure for Time Series. Physical Review Letters. 2002;88(17):174102.

25. Richman JS, Moorman JR. Physiological time-series analysis using approximate entropy and sample entropy. American Journal of Physiology-Heart and Circulatory Physiology. 2000;278(6):H2039–H49.

26. Struys MM. Spectral Entropy as an Electroencephalographic Measure of Anesthetic Drug Effect: A Comparison with Bispectral Index and Processed Midlatency Auditory Evoked Response. Anesthesiology. 2004;101(1):34–42.

27. Zemp R, Tanadini M, Plüss S, Schneller M, Meboldt M, Taylor WR, et al. Application of Machine Learning Approaches for Classifying Sitting Posture Based on Force and Acceleration Sensors. BioMed Research International. 2016.

28. Kuhner A, Schubert T, Cenciarini M, et al. Correlations between Motor Symptoms across Different Motor Tasks, Quantified via Random Forest Feature Classification in Parkinson’s Disease. Frontiers in Neurology. 2017;8:607.

29. Pavey TG, Gilson ND, Gomersall SR, Clark B, Trost SG. Field evaluation of a random forest activity classifier for wrist-worn accelerometer data. Journal of Science and Medicine in Sport. 2017;20(1):75–80.

30. Díaz-Uriarte R, de Andrés SA. Gene selection and classification of microarray data using random forest. BMC Bioinformatics. 2006;7:3.

31. Li B-Q, Cai Y-D, Feng K-Y, Zhao G-J. Prediction of Protein Cleavage Site with Feature Selection by Random Forest. PLoS ONE. 2012;7(9).

32. Mursalin M, Zhang Y, Chen Y, Chawla NV. Automated epileptic seizure detection using improved correlation-based feature selection with random forest classifier. Neurocomputing. 2017;241:204–14.

33. Breiman L. Random Forests. Machine Learning. 2001;45(1):5–32.

34. Vanmechelen I, Bekteshi S, Bossier K, Feys H, Deklerck J, Monbaliu E. Presence and severity of dystonia and choreoathetosis overflow movements in participants with dyskinetic cerebral palsy and their relation with functional classification scales. Disability and rehabilitation. 2019:1.

35. Al AS, editor Dyskinesia and motor state detection in Parkinson’s Disease patients with a single movement sensor. 2012 Annual International Conference of the IEEE Engineering in Medicine and Biology Society; 2012; San Diego, CA.

36. Keijsers NLW, Horstink MWIM, Gielen SCAM. Automatic assessment of levodopa-induced dyskinesias in daily life by neural networks. Movement Disorders. 2003;18(1):70–80.

37. Patel S, Lorincz K, Hughes R, et al. Monitoring motor fluctuations in patients with Parkinson’s disease using wearable sensors. IEEE Transactions on Information Technology in Biomedicine. 2009;13:864–73.

38. den Hartog D, van der Krogt MM, van der Burg S, Aleo I, Gijsbers J, Bonouvrié LA, et al. Home-Based Measurements of Dystonia in Cerebral Palsy Using Smartphone-Coupled Inertial Sensor Technology and Machine Learning: A Proof-of-Concept Study. Sensors. 2022;22(12):4386.

39. Haberfehlner H, Roth Z, Vanmechelen I, Buizer AI, Vermeulen RJ, Koy A, et al. A Novel Video-Based Methodology for Automated Classification of Dystonia and Choreoathetosis in Dyskinetic Cerebral Palsy During a Lower Extremity Task. Neurorehabilitation and neural repair. 2024;38(7):479–92.

40. Vanmechelen I, Danielsson A, Lidbeck C, Tedroff K, Monbaliu E, Krumlinde-Sundholm L. The Dyskinesia Impairment Scale, Second Edition: Development, construct validity, and reliability. Developmental Medicine & Child Neurology. 2023;65(5):683–90.

